# Making densenet interpretable a case study in clinical radiology

**DOI:** 10.1101/19013730

**Authors:** Kwun Ho Ngan, Artur d’Avila Garcez, Karen M. Knapp, Andy Appelboam, Constantino Carlos Reyes-Aldasoro

## Abstract

The monotonous routine of medical image analysis under tight time constraints has always led to work fatigue for many medical practitioners. Medical image interpretation can be error-prone and this can increase the risk of an incorrect procedure being recommended. While the advancement of complex deep learning models has achieved performance beyond human capability in some computer vision tasks, widespread adoption in the medical field has been held back, among other factors, by poor model interpretability and a lack of high-quality labelled data. This paper introduces a model interpretation and visualisation framework for the analysis of the feature extraction process of a deep convolutional neural network and applies it to abnormality detection using the musculoskeletal radiograph dataset (MURA, Stanford). The proposed framework provides a mechanism for interpreting DenseNet deep learning architectures. It aims to provide a deeper insight about the paths of feature generation and reasoning within a DenseNet architecture. When evaluated on MURA at abnormality detection tasks, the model interpretation framework has been shown capable of identifying limitations in the reasoning of a DenseNet architecture applied to radiography, which can in turn be ameliorated through model interpretation and visualization.

## 1. INTRODUCTION

Radiography is a medical imaging technique that uses X-rays to generate images of human anatomy [1]. X-rays have now been used for more than one hundred years for the different applications like detection of fractures [2] or detection of obstruction of the duodenum [3]. The rays penetrate the object of interest, say a wrist or an elbow, and are attenuated according to the density of the material they travel through. Thus denser materials, such as bones, present a higher attenuation than softer materials, such as muscles or organs. The density is normally indicated in images as various levels brightness (dense) or darkness (soft), although in some cases the scale is reversed. It is important to notice that X-ray images are two dimensional projections of three dimensional bodies, and thus in some cases two or more images are acquired with different angles of view like posterior-anterior (e.g. Figure 1(a)) or lateral views (e.g. Figure 1(b)). Metallic objects are denser than bones and appear much brighter than bones.

**Fig. 1.**
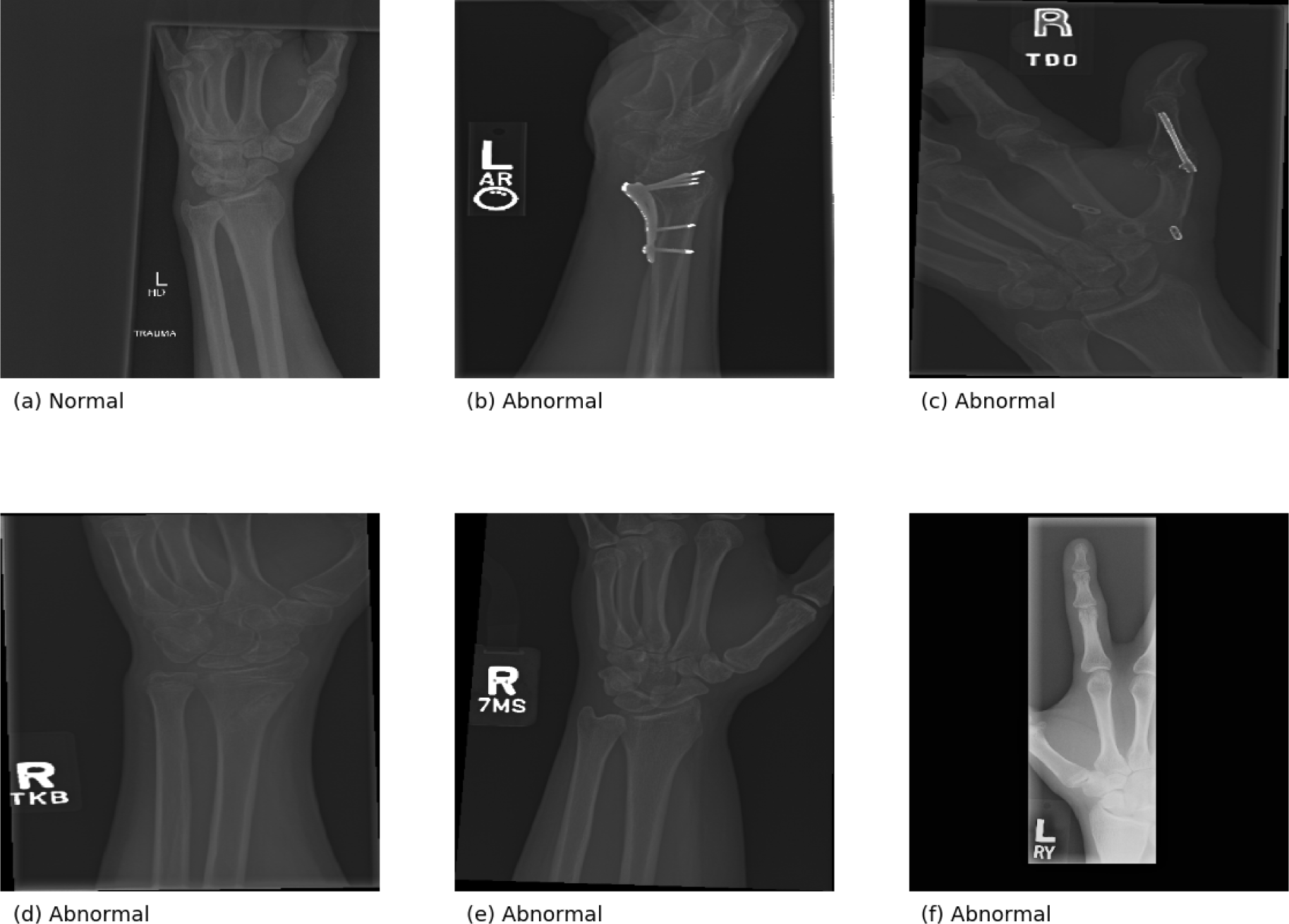
Representative images from the MURA dataset [10]. (a) Normal wrist. (b) Foreign objects at the wrist. (c) Foreign objects in thumb. (d) Wrist fracture. (e) Degenerated wrist joint. (f) Degenerated finger joint.

Radiographic images are generally analyzed by certified radiologists or specialist reporting radiologists. On a typical day, a single analysis is performed under a tight time constraint alongside other heavy workloads. It has been reported that work fatigue can lead to errors in interpretation [4]. Therefore, it is attractive to explore computational approaches that can support radiologists in the analysis of X-ray images.

Recent advances in the areas of machine learning, computer vision and especially deep learning model development have reported results where the computational models have surpassed human performance on many predictive tasks. In 2012, the Alexnet [5] won the ImageNet Large Scale Visual Recognition Competition (ILSVRC) by a large margin against the runner-up. More sophisticated approaches have since improved the accuracy obtained with Alexnet. ResNet [6] developed by Microsoft, has outperformed human prediction in the same competition by 2015.

Despite their superior performance, widespread adoption of these complex deep learning models has been held back primarily due to poor model interpretability, the requirement of a very large number of annotated cases to train the models and of sophisticated computational resources. The lack of model transparency of its predictions, in particular, has led to challenges in obtaining regulatory approval to deploy in life-critical applications for healthcare.

Previous work has described the association between key features and the predicted classes [7, 8, 9]. While it is important to learn about these attributing features, it is equally important to understand the feature extraction mechanism of a deep learning model when our goal is to optimise the model design and its training regime. Being able to understand and describe the feature extraction process will also facilitate reasoning about the model and model explanation to other stakeholders (e.g. clinicians or radiologists) who might be more interested in the attributes of the original data rather than the models themselves.

This paper introduces a systematic framework to explore the feature extraction mechanism in a deep learning model applied to images. The framework is evaluated on a DenseNet model [10, 11] by producing a combination of model visualisation and reasoning from layer-wise and class activations. An initial analysis from expert radiologists has also been carried out.

The contribution of this paper is two-fold: (1) The proposed framework allows a systematic approach for analysing the feature extraction mechanism in the DenseNet model; (2) The effectiveness of the approach is evaluated using MURA, showing that the framework is capable of identifying and mitigating certain hidden feature extraction failures through a visually explainable manner.

The rest of the paper is organised as follows. Section 2 reviews the relevant literature that has shaped the framework, from the choice of dataset and network architecture to the methods of visualisation. Section 3 introduces the model interpretation framework with emphasis on how the model is trained and then used for layer-wise interpretation and reasoning about its predictions. Experimental results are reported in Section 4) and further discussed from a clinical perspective in Section 5. Section 6 concludes and discusses directions for future work.

## 2. BACKGROUND AND RELATED WORK

The architecture of Dense Convolution Networks (DenseNet) was developed by [11]. It has incorporated several promising improvements over previous deep learning models such as the alleviation of vanishing gradient, strengthening of feature propagation and better feature reuse while substantially reducing the total number of model parameters in comparison with similar models like ResNet [6]. Figure 2 presents a schematic representation of the DenseNet architecture for an end-to-end prediction using input from a medical image.

**Fig. 2.**
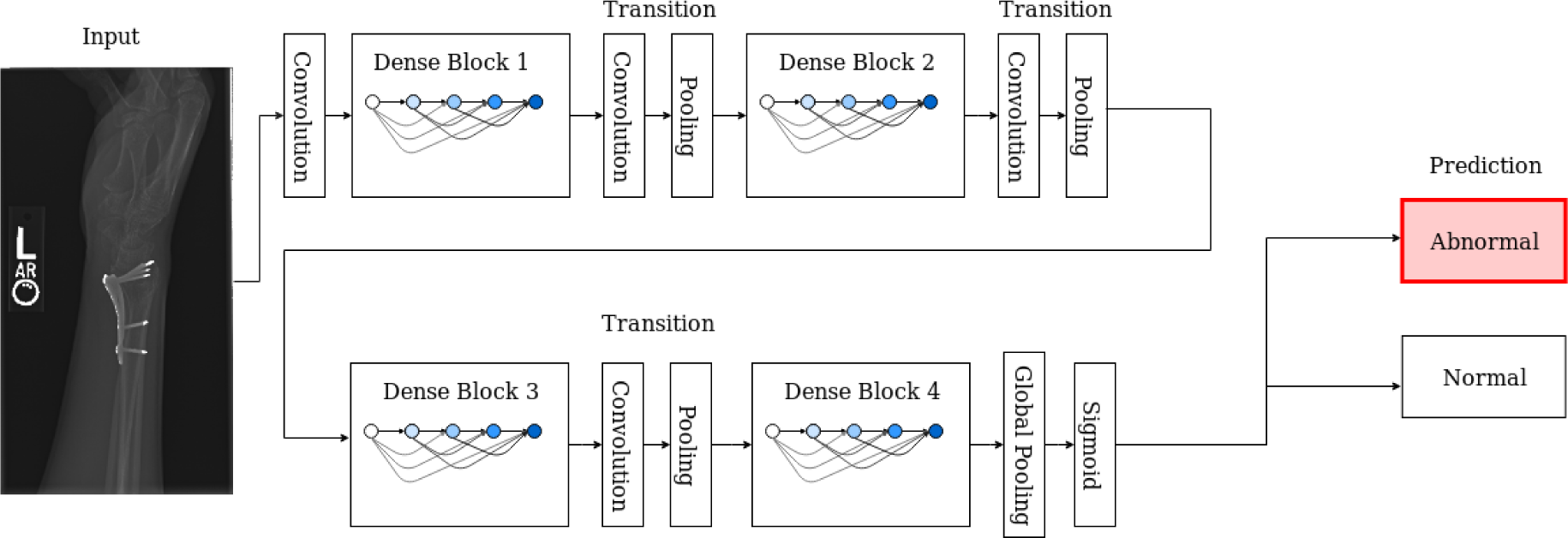
A schematic representation of the 169-layered DenseNet architecture with four dense blocks. The transition layers between blocks formed by convolution and pooling are used for down-scaling the feature maps.

In a DenseNet architecture, a common composite function was applied in blocks for non-linear transformation. This composite function was made up of a series of consecutive operations including Batch Normalisation (BN), Activation with Rectified Linear Unit (ReLU) and a 3⨯3 convolution (Conv). To enhance computational efficiency, a bottleneck layer of 1⨯1 convolution was also introduced in the original work to reduce the number of feature maps generated. Concatenating layers were applied to capture information from feature maps of all preceding layers within each dense block. Transition layers were implemented between blocks to consolidate the feature maps from the previous block and transpose them into down-scaled feature maps for the next block. The number of feature maps (i.e. the amount of new information allowed for extraction) is regulated by a user-defined global parameter (i.e. growth rate).

[10] compiled one of the largest public collections (MURA) for de-identified, Health Insurance Portability and Accountability Act (HIPAA)-compliant musculoskeletal radiographic images. The images are sourced from 12,173 patients with 14,863 studies (a total of 40,561 images captured from multiple angles). These images are categorised into seven extremity types (elbow, finger, forearm, hand, humerus, shoulder and wrist). The dataset has been split into training and validation by the data maintainers. Each study is labelled by certified radiologists with an average of 8.83 years of experience. The studies can either be classified as negative (normal) or positive (abnormal) according to the presence of abnormalities such as fracture, hardware implant, degenerative joint diseases or other causes (e.g. lesions and subluxations). Figure 1 presents sample images from the dataset that will be used in this work.

In the original work of [10], a 169-layered DenseNet architecture was used as a model for performance evaluation. It was reported that the performance of the model varied significantly across different extremity types. On wrist cases the model performed well, even exceeding human prediction. On other studies the model had failed to achieve similar accuracy. In the case of fingers, both human and model were unable to predict well against the ground truth.

In the work of [11] and [10], their models were pre-trained using the ImageNet dataset and the model weights were refined during a transfer learning stage. ImageNet [12] is a large hierarchical database of human-annotated images intended for general-purpose computer vision research. At present, it consists of approximately 14 million images in more than 21 thousand classes (synsets). [13] has conducted an empirical investigation on the performance of applying ImageNet in transfer learning. It was reported that the pre-trained weights from the ImageNet dataset could significantly enhance feature extraction even when the model was trained from images unrelated to the target task. Huh’s work [13] had also demonstrated the impact from image quantity, class size and level of visual similarity among images on transfer learning providing suggestions to some of our investigations presented in this work.

The performance of deep learning models has been evaluated extensively through various statistical measures. The interpretation of these models has however only begun in recent years through model visualisation [14, 15] and feature association with prediction [8, 9]. In particular, the layer-wise activations shown in [14] and class activation maps from [15] have inspired our proposed framework to understand how relevant features are extracted across a deep learning network and a mean for human explanation.

## 3. PROPOSED FRAMEWORK

This work applied a 169-layered DenseNet architecture as a trained model for model visualisation and interpretation. The model was constructed using Keras/Tensorflow under a Linux based environment. The initial weights of the network were either randomly assigned or loaded with weights pre-trained using the ImageNet dataset. In both cases, transfer learning was completed by an end-to-end re-training of the models with 32 epochs using training data from the MURA dataset.

During model re-training, four sets of model weights had been generated, namely, (1) a pre-trained model with training data of all extremity types (2) a pre-trained model with training data of wrist-specific images (3) a randomly initialised model with training data of wrist-specific images, and (4) a pre-trained model with training data of finger-specific images. Most of the model hyper-parameters used in this work followed closely to the defined setup in both [10] and [11]’s work. Some minor variations had been implemented for simpler execution. The model was trained end-to-end in minibatches of size 8. An initial learning rate of 0.0001 was used with a decay factor of 10, 50 and 100 respectively when 25%, 50% and 75% of training epochs have lapsed. A sigmoid function was used in the last layer for binary classification. During inference, a final prediction value above 0.5 was regarded as positive (i.e. abnormal) and negative (normal) otherwise. A loss function of binary cross-entropy was optimised using Adam with default parameters *β*^1^ being 0.9 and *β*^2^ being 0.999 [16]. The best model in each model set was selected among all training epochs for which the difference between training and validation accuracy was the least to avoid over-fitting. All the best models were then used for model evaluation and interpretation throughout this work.

Input images for the models were pre-processed by scaling to a standard size of 320 ⨯320. As the original DenseNet model was trained with an input size of 224 × 224 [11], the pre-trained weights had to be modified through Keras’ built-in conversion function. Images were also randomly augmented during training with random lateral inversions and rotations up to 30 degrees.

The models were primarily evaluated using standard classification accuracy and Cohen’s Kappa statistics at the individual image level. This differs from the Cohen’s Kappa statistics at the study level presented in [10] which measures an aggregated prediction based on the majority vote from the classification of multiple images in the same study. Thus, Table 1 should not be a direct comparison with the results of [10]. The authors believe that the performance metrics at image level provides better model interpretation comparison. It would also permit a more consistent performance comparison for using a similar setup of the DenseNet architecture on other image datasets in the future.

**Table 1.** Performance metrics for the different model sets on an individual image basis

| Model Set | Validation Accuracy | Cohen’s Kappa Statistics |
| --- | --- | --- |
| All Extremity Types (pre-trained) | 0.830 | 0.658 |
| Wrist-Only (pre-trained) | 0.862 | 0.717 |
| Wrist-Only (random initialised) | 0.778 | 0.539 |
| Finger-Only (pre-trained) | 0.766 | 0.622 |

**Table 2.**
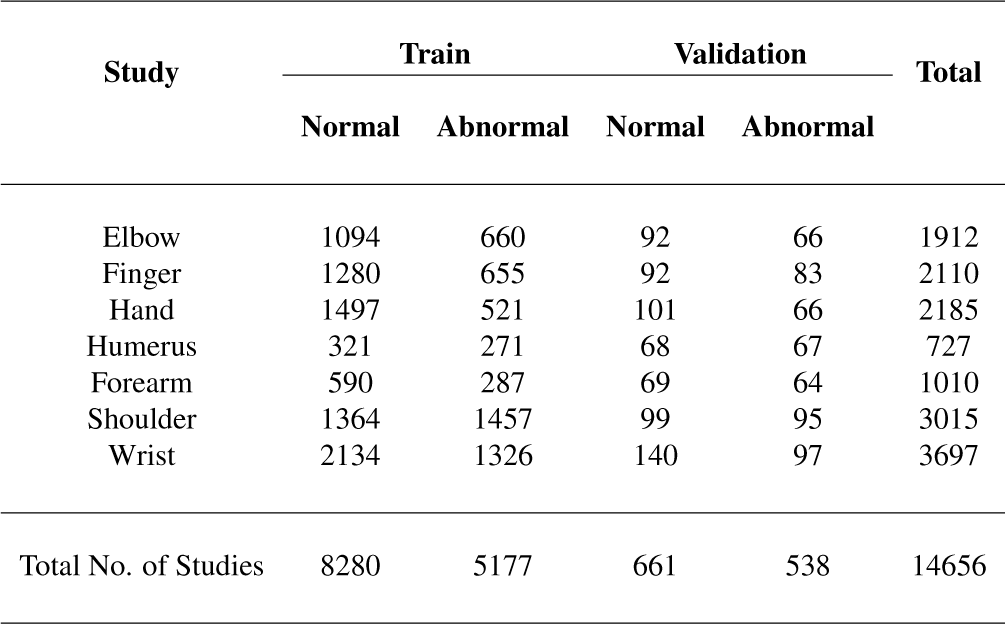
The distribution of studies in the MURA dataset with 9,045 normal and 5,818 abnormal cases [10]

In order to interpret the model feature representations, visualisation of layer-wise activations was plotted using Matplotlib with a ‘bwr’ colour map (blue, white, red). The range of activation values was represented by the colour of blue for the low spectrum (i.e. near-zero activations) and red for the significant features that would be passed on to the subsequent layers. Class activation map (CAM) [15] was also applied to the final convolution layer before the sigmoid function. This class activation map used a jet colour map (blue to red through the rainbow colours). The map was upscaled and overlayed over the original input image to visualise the final extracted features contributing to the abnormality detection. A schematic representation for the proposed framework during prediction is shown in Figure 3.

**Fig. 3.**
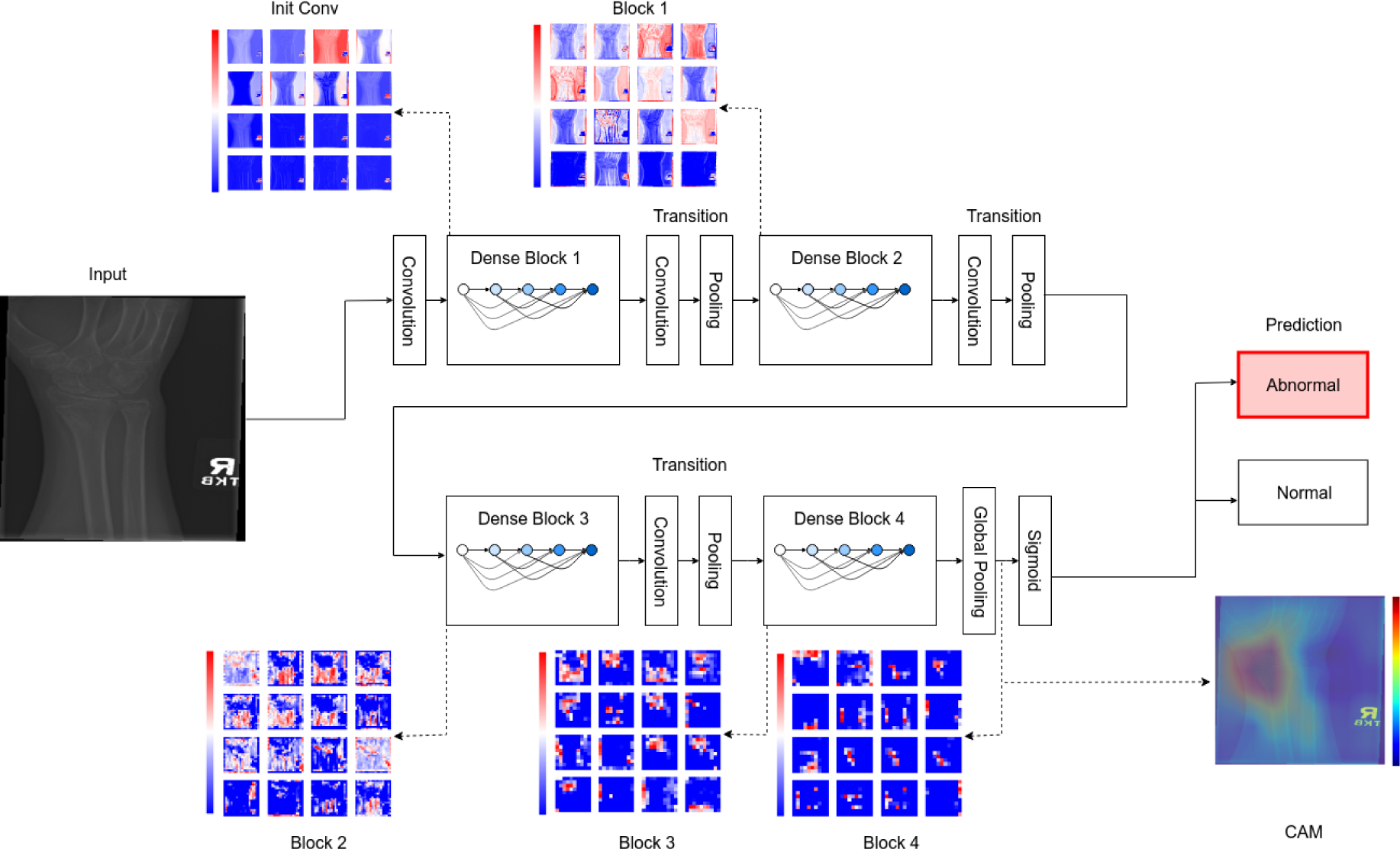
Schematic representation of an X-ray image abnormality detection and model interpretation using layer-wise and class activation maps across different dense blocks. At each level, a series of feature maps are generated, the resolution decreases with the progress through the blocks. Colours are mapped to represent the range of activation values with blue in the lower spectrum for near-zero activation and red for the highly activated features. The final output, labelled here as CAM, is a feature map that highlights the area(s) where abnormalities can be located. The regions with warm colours indicate higher probability than the areas with cool colours.

## 4. EXPERIMENTAL RESULTS

The first model evaluation was conducted through the performance comparison from the best selected models on validation data of the corresponding extremity types (e.g. wrist-only, finger-only, etc.). From the values presented in Table 1, all models have acquired relevant learning during model training to a varying degree. All models achieved an accuracy of approximately 80% without any attempt of model optimization. Interestingly, the wrist-only model appears to detect abnormality on wrist better than a finger-only model on cases of fingers. The all-extremity model used in this paper was also found comparable with similar DenseNet single models in the MURA leaderboard on a study level basis as shown in Table 3 & 4 where this model was evaluated based on the average Cohen’s Kappa score of 100 randomly selected sample set from the validation image collection.

**Table 3.** Leaderboard for MURA Dataset [10]

| Rank | Date | Model | Kappa |
| --- | --- | --- | --- |
| 1 | Nov 20, 2018 | base-comb2-xuan-v3(ensemble) jzhang Availink | 0.843 |
| 2 | Nov 06, 2018 | base-comb2-xuan(ensemble) jtzhang Availink | 0.834 |
| 3 | Oct 06, 2018 | multi.type (ensemble model) SCU_MILAB | 0.833 |
| 4 | Oct 02, 2018 | base-comb4(ensemble) jtzhang Availink | 0.824 |
| 5 | Nov 08, 2018 | base-comb2-jun2(ensemble) | 0.814 |
| 5 | Nov 07, 2018 | base-comb2-ping(ensemble) | 0.814 |
| 6 | Aug 22, 2018 | base-comb3(ensemble) | 0.805 |
| 7 | Sep 14, 2018 | double.res(ensemble model) SCU_MILAB | 0.804 |
| 8 | Aug 24, 2018 | double-dense-Axy-Axyf512 ensemble | 0.795 |
| 9 | Jul 24, 2018 | he-j | 0.775 |
| 10 | Aug 19, 2018 | ianpan (ensemble) RIH 3D Lab | 0.774 |
| 11 | Jul 24, 2018 | he-j | 0.774 |
| 12 | Jun 17, 2018 | gcm (ensemble) Peking University | 0.773 |
| 12 | Sep 10, 2018 | ty101 single model | 0.773 |
| 13 | Aug 31, 2018 | he-j | 0.764 |
| 13 | Aug 31, 2018 | AIAPlus (ensemble) Taiwan AI Academy | 0.764 |
| 14 | Sep 04, 2018 | SER_Net.Baseline (single model) SJTU | 0.764 |
| 15 | Jul 14, 2018 | Trs (single model) SCU_MILAB | 0.763 |
| 16 | Sep 12, 2018 | null | 0.763 |
| 16 | Sep 12, 2018 | ellonde | 0.763 |
| 17 | Jul 16, 2018 | null | 0.755 |
| 17 | Aug 24, 2018 | dense-sep-xyz ensemble | 0.755 |
| 18 | Nov 16, 2018 | VGG19 single model | 0.754 |
| 19 | Jul 25, 2018 | DenseNet001 (single model) zhou | 0.747 |
| 20 | Aug 21, 2018 | dn169-Aftrva(single) AliHealth | 0.747 |
| 21 | Jul 14, 2018 | type_resnet (single model) CCLab | 0.746 |
| 22 | Dec 06, 2018 | res101_da_sqtv(single) | 0.746 |
| 23 | Jun 18, 2018 | VGG19 (single model) ZHAW | 0.744 |
| 24 | Jul 02, 2018 | ImageXrModel-001 (single model) Ruslan Baikulov | 0.737 |
| 25 | Oct 04, 2018 | ExtremityModel ensemble | 0.734 |
| 26 | Jan 19, 2019 | DenseAttention (single model) BIT | 0.727 |
| 26 | Aug 12, 2018 | base169-AllParts-diffParts-tv(ensemble) MSFT-research | 0.727 |
| 27 | Sep 27, 2018 | aiinside | 0.725 |
| 27 | Mar 14, 2019 | Resology14 (ensemble) Rology | 0.725 |
| 28 | Dec 06, 2018 | inc3_sqtv(single) MIT AI | 0.724 |
| 29 | Aug 21, 2018 | base-model-Atv(single) Avail-AI | 0.717 |
| 30 | Dec 11, 2018 | incev3_xy(single) UCB | 0.716 |
| 31 | Jul 18, 2018 | he-j | 0.712 |
| 32 | Mar 14, 2019 | kmle-second (ensemble) kmle | 0.707 |
| 33 | Jul 30, 2018 | he-j | 0.712 |
| 34 | May 23, 2018 | Stanford Baseline (ensemble) Stanford University | 0.705 |
| 35 | Mar 10, 2019 | asa_model_nasnetmo (single) toyohashi | 0.702 |
| 36 | Jul 10, 2018 | he-j | 0.701 |
| 36 | Jul 16, 2018 | type_inception2(single model) CCLab | 0.701 |
| 37 | Dec 06, 2018 | term2-model0sqtv(single) | 0.700 |
| 38 | Jun 24, 2018 | single-densenet169 single model | 0.699 |
| 39 | Oct 26, 2018 | Joint-tv single | 0.698 |
| 39 | Aug 17, 2018 | base ensemble | 0.698 |
| 39 | Aug 21, 2018 | baseAllPartsDiffParts-sq ensemble | 0.698 |
| 39 | Aug 09, 2018 | base169-AllParts-diffParts(ensemble) MSFT-research | 0.698 |
| 39 | Jul 02, 2018 | Baseline169 (single model) Personal | 0.698 |
| 40 | Jan 18, 2019 | first-attempt-kmle (ensemble) kmle | 0.696 |
| 41 | Dec 31, 2018 | DenseNet_144 single model | 0.694 |
| 42 | Jul 22, 2018 | Densenet DI-MT Single | 0.690 |
| 43 | Jul 13, 2018 | dense169(ensemble) mitAI | 0.686 |
| 44 | Oct 27, 2018 | xception(single model) bimal | 0.686 |
| 45 | Dec 23, 2018 | {EnglebertDGC} (single model) UCLouvain | 0.684 |
|  |  | **DenseNet169 (single model) This Paper** | **0.683** |
| 46 | Dec 06, 2018 | res_daxy(single) CMU ml | 0.680 |
| 47 | Jan 16, 2019 | GoGoing (ensemble) Inner Mongolia University | 0.678 |
| 48 | Dec 11, 2018 | inceptionresnetv2.tv(single) baidu AI | 0.676 |
| 49 | Jun 10, 2018 | ResNet (single model) UCSC CE graduate student huimin yan | 0.675 |
| 49 | Aug 18, 2018 | {monica_v1}(single model) Zzmonica | 0.675 |
| 50 | Jul 09, 2018 | null | 0.664 |
| 51 | Jan 11, 2019 | PfResNet (single model) USTC_Math_1222 | 0.664 |
| 52 | Nov 29, 2018 | {DenseNet_169} (single model) Rology | 0.662 |
| 53 | Jan 31, 2019 | DenseNet_v2 single model | 0.661 |
| 54 | Sep 03, 2018 | DenseNet002 (single model) zhou | 0.660 |
| 55 | Nov 05, 2018 | DenseNet (single model) Rology | 0.659 |
| 56 | Jun 30, 2018 | Baseline169-v0.2 (single) Personal | 0.659 |
| 57 | Jul 08, 2018 | madcarrot | 0.653 |
| 58 | Dec 06, 2018 | base_largexy(single) Tsinghua Deep Learning | 0.652 |
| 59 | Jun 30, 2018 | zhy | 0.638 |
| 60 | Jun 30, 2018 | Densenet121 (single model) Personal | 0.629 |
| 61 | Oct 26, 2018 | baseJoint-tvsq(single) ali | 0.624 |
| 62 | Dec 31, 2018 | ConvNet single model | 0.599 |
| 63 | Jan 31, 2019 | Ensemble_V0 ensemble model | 0.599 |
| 64 | Aug 29, 2018 | Inception-ResNet-v2 (single model) Royal Holloway | 0.597 |
| 64 | Aug 28, 2018 | Inception-ResNet-v2 (single model) Royal Holloway | 0.597 |
| 65 | Jul 26, 2018 | Bhaukali_v1.0 (single model) IIT BHU, Varanasi | 0.581 |
| 66 | Jul 21, 2018 | inceptionv3-pci (single model) PCI | 0.578 |
| 67 | Jul 12, 2018 | DN169 single | 0.574 |
| 68 | Jul 31, 2018 | Densenet169-lite(single model) Tang | 0.560 |
| 69 | Aug 29, 2018 | ensemble1 ensemble | 0.534 |
| 70 | Jul 06, 2018 | DenseNet (single model) Zhou | 0.518 |

**Table 4.** Comparative Performance for Single DenseNet model on a study-level basis [10]

| Rank | Date | Model | Kappa |
| --- | --- | --- | --- |
| 19 | Jul 25, 2018 | DenseNet001 (single model) zhou | 0.747 |
| 20 | Aug 21, 2018 | dn169-Aftrva(single) AliHealth | 0.747 |
| 26 | Jan 19, 2019 | DenseAttention (single model) BIT | 0.727 |
| 38 | Jun 24, 2018 | single-densenet169 single model | 0.699 |
| 39 | Jul 02, 2018 | Baseline169 (single model) Personal | 0.698 |
| 41 | Dec 31, 2018 | DenseNet_144 single model | 0.694 |
| 42 | Jul 22, 2018 | Densenet DI-MT Single | 0.690 |
|  |  | **DenseNet169 (single model) This Paper** | **0.683** |
| 52 | Nov 29, 2018 | {DenseNet_169} (single model) Rology | 0.662 |
| 53 | Jan 31, 2019 | DenseNet_v2 single model | 0.661 |
| 54 | Sep 03, 2018 | DenseNet002 (single model) zhou | 0.660 |
| 55 | Nov 05, 2018 | DenseNet (single model) Rology | 0.659 |
| 56 | Jun 30, 2018 | Baseline169-v0.2 (single) Personal | 0.659 |
| 60 | Jun 30, 2018 | Densenet121 (single model) Personal | 0.629 |
| 67 | Jul 12, 2018 | DN169 single | 0.574 |
| 68 | Jul 31, 2018 | Densenet169-lite(single model) Tang | 0.560 |
| 70 | Jul 06, 2018 | DenseNet (single model) Zhou | 0.518 |

In order to investigate the contributing factors for the model performance, layer-wise and class activations were applied on selected images to understand the feature extraction mechanism performed by the DenseNet architecture. Figure 4 presents an example of activation visualisations that will be used throughout this work for analysis and comparison. All other selected cases can be found in Appendix A.2. It should be noted that the presented cases in this work may not capture all scenarios in the dataset. However, the authors believe these cases are representative and sufficient to demonstrate the effectiveness of the proposed framework for the model interpretation purpose.

**Fig. 4.**
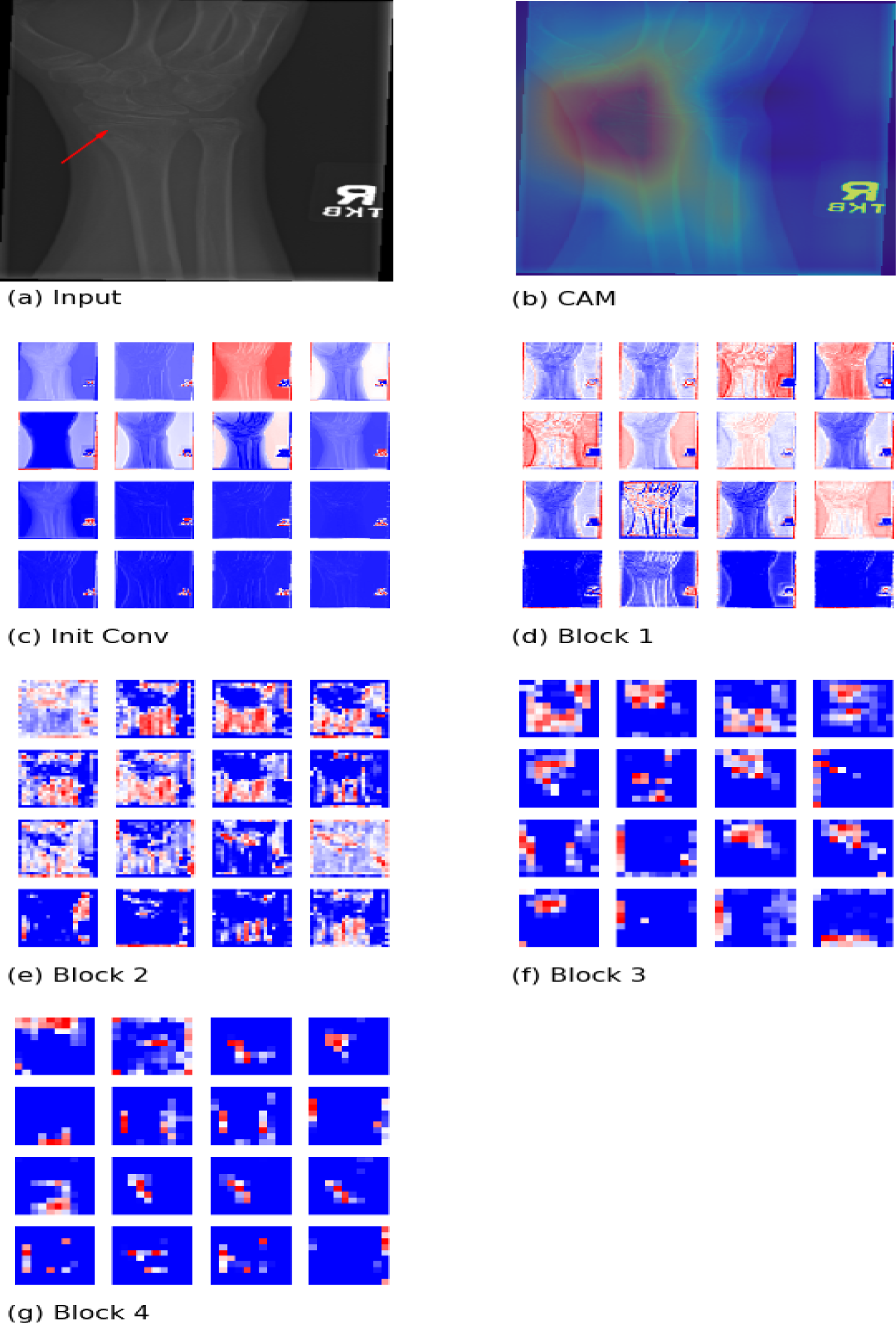
Case 1: Feature extraction for a case of wrist fracture using a wrist-only pre-trained model for the radiograph shown in Fig. 1(d). The arrow indicates a fracture. The extraction began with an augmented input image (a). High-level features (e.g. foreground extraction and regions of the wrist) were extracted in the earlier blocks and can be seen from the corresponding activation maps (c, d, e). Fine details (e.g. location of fracture) were extracted in block 3 and 4 (f, g). A class activation map at the last layer is presented in (b) where the final attributing features were identified, highlighting the area of interest.

Figure 4 shows a case of wrist fracture as highlighted by the red arrow in Figure 4a. When an augmented image was inputted into the model, the top 16 most activated feature maps were generated for the chosen activation layers at the end of each stage across different dense blocks. The final feature maps (Figure 4g) presented the same features that made up the class activation map.

From the figure, it can be found that the feature mechanism followed an intuitive hierarchical path. The model began by identifying the foreground in the initial convolution (Figure 4c). The region of the wrist could be outlined by the end of block 2 (Figure 4e), and fine-grained features were extracted in block 3 and 4 with increasing precision. At the end of the network, most of the feature maps were activated close to where the fracture was located. It should however be noted that the resolution at the final activation layer was significantly coarser than the input image. These visualisations should therefore be regarded as a mean for understanding the path of feature extraction. Precise localisation of abnormality will be beyond the scope of this work.

Among the cases studied in Appendix A.2, hardware implants were found to be more easily detectable as abnormality while degenerative joints appeared misclassified as normal by the model. For example in Figure 5, the screw fixation and the text legends were detected before the end of block 1. Within block 3 and 4 (Figures 5f, 5g), the model attempted to refine the separation between the fixation and the text legends. Although many final feature maps in block 4 captured only the fixation, the text legend remained a more significant feature in the class activation map making the prediction being biased to the wrong feature. In fact, the prediction of this image should be regarded as misclassified given the incorrect attributing features for abnormality. This is a type of error that is very difficult to detect without performing a similar framework of model visualisation presented in this work.

**Fig. 5.**
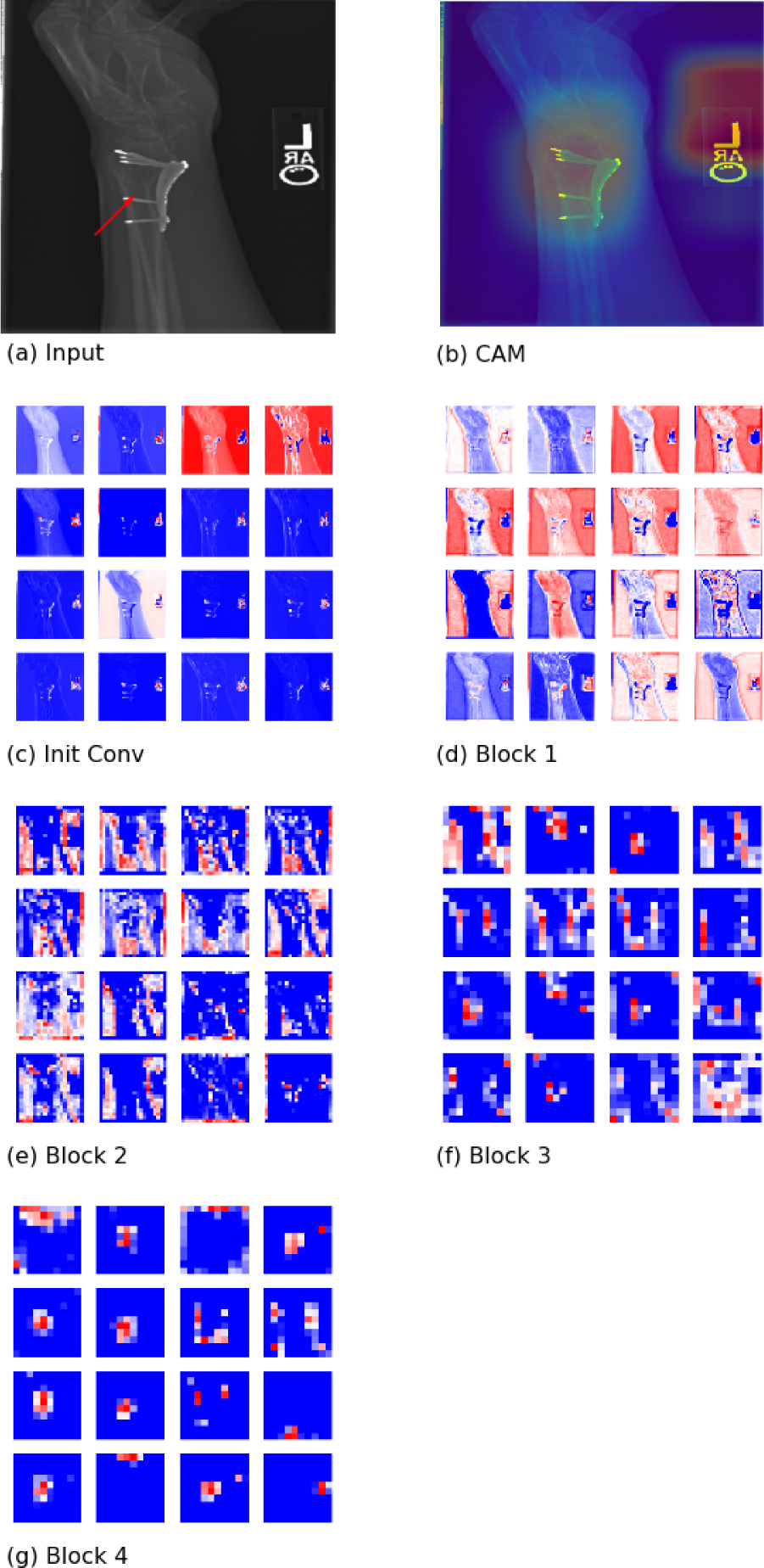
Feature extraction from an image with screw fixation at the wrist. The fixation could be identified as early as the initial convolution (c). However, the fixation was always coupled with the text legend in all feature maps. Separation of these two objects was attempted in subsequent blocks (d - g). It can be observed that only the screw fixation was found in some feature map by the end of block 4 (g). Nevertheless, the text legend remained a key feature contributing to the abnormality detection in the class activation map (b).

Degenerative joint diseases are complicated to analyse even from human eyes. Figure 6 presents a false negative case where there is a shifted scaphoid bone at the wrist joint. This image was classified by the model as normal with a blue region around the wrist in class activation. All other regions are regarded as abnormal in relative terms. By looking at the layer-wise activations, the feature extraction mechanism remained the same. Nevertheless, it seemed that the model did not have the capacity in block 3 and 4 to differentiate this abnormality from the bone alignment of a normal wrist.

**Fig. 6.**
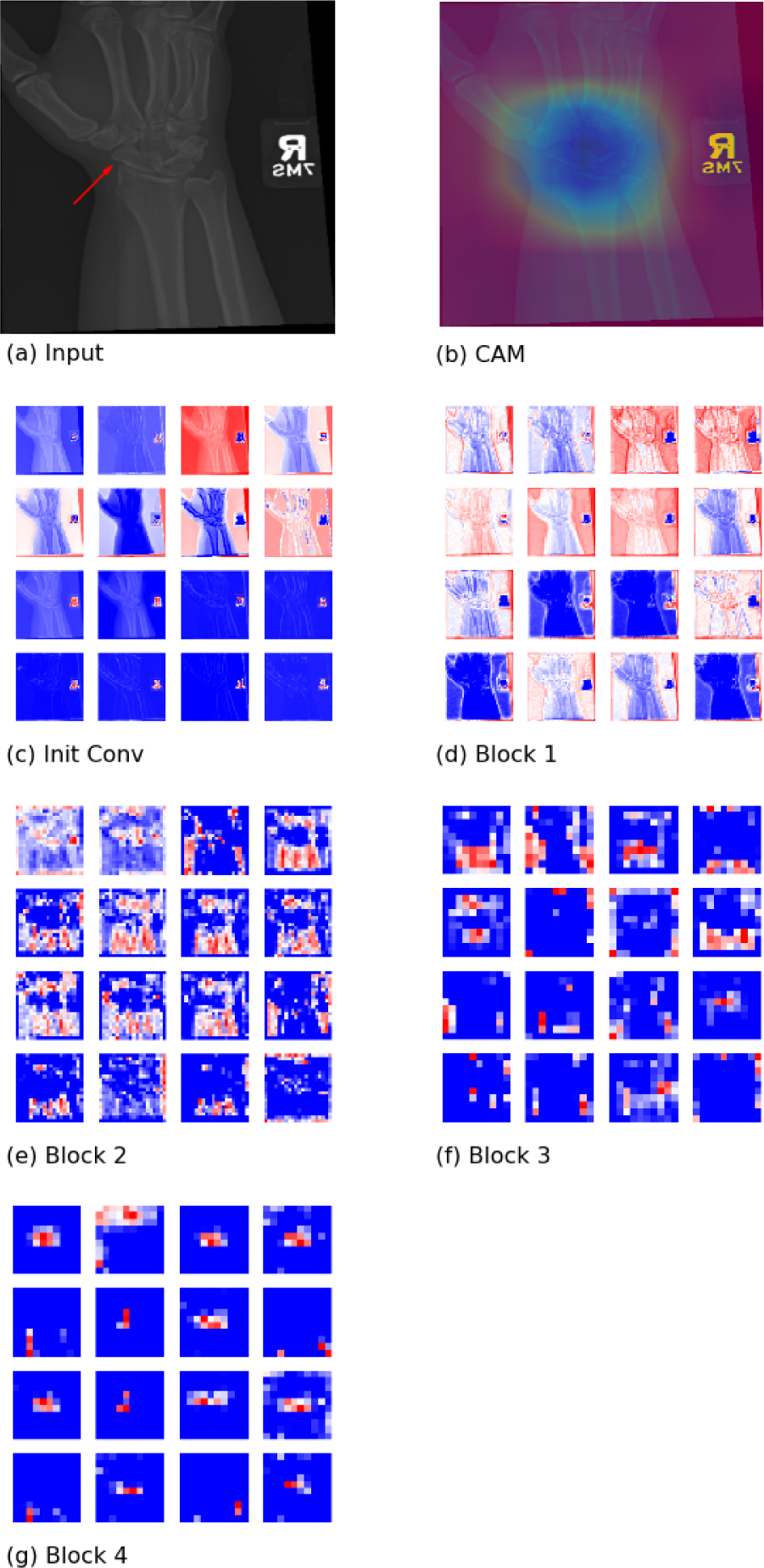
Feature extraction from an image with shifted scaphoid bone at the wrist joint. The foreground and the region of wrist were extracted in the earlier blocks (c, d & e). Fine-grained feature extraction were attempted in block 3 and 4 (f & g). However, the model capacity in these blocks might not be sufficient to capture the details of the bone shift.

This framework of model interpretation was also extended to investigate the effect of training image quantity. It can be observed in Figure 7 that the use of pre-trained weight from ImageNet was highly valuable. Without it, the overall structure of the wrist could not be identified in the earlier layers. This had led to challenges in breaking down the image into relevant regions and forced the model to identify abnormality from within the entire middle region (i.e. the blue region in Figure 7f) of the image. It is also interesting to note that the extraction of the text legend remained highly effective. This may be somewhat expected given that the text legends were present in most of the training images within the MURA dataset. Furthermore, a pre-trained model with training data from all extremity types (i.e. even more images with text legends) was found to minimise the bias of text legend in the case of screw fixation (Figure 8). The final features at the end of Block 4 (Figure 8g) were primarily pixels around the fixation. The investigations made in cases of fingers had found similar success. Nails implanted in a thumb could be identified by the end of Block 1 (Figure 9d). Sufficient capacity within the subsequent layers allowed for excellent refinement of the key features extracted. In Figure 10, a highly challenging case was presented where there was an extra rectangular frame commonly found in images of the MURA dataset. The challenge was also coupled with a task to predict a misaligned fingertip. From the layer-wise activations, it can be noticed that the model was not able to identify the hand and fingers in the early layers (i.e. before the end of block 1). The influence of the frame had propagated across the entire network. The border of the frame eventually formed a significant portion of the final extracted features by the end of block 4 (Figure 10g). The issue of image frame could easily be mitigated with simple image cropping. A case using the cropped image is presented in Figure 11. The identification of the hand and finger had been significantly improved. The only remaining issue was the insufficient learning within the model to differentiate a degenerated joint or deformation. Improving the learning in this aspect will potentially improve the accuracy in cases of fingers as degenerated joint disease and deformation are more commonly found as abnormality in X-ray images of fingers.

**Fig. 7.**
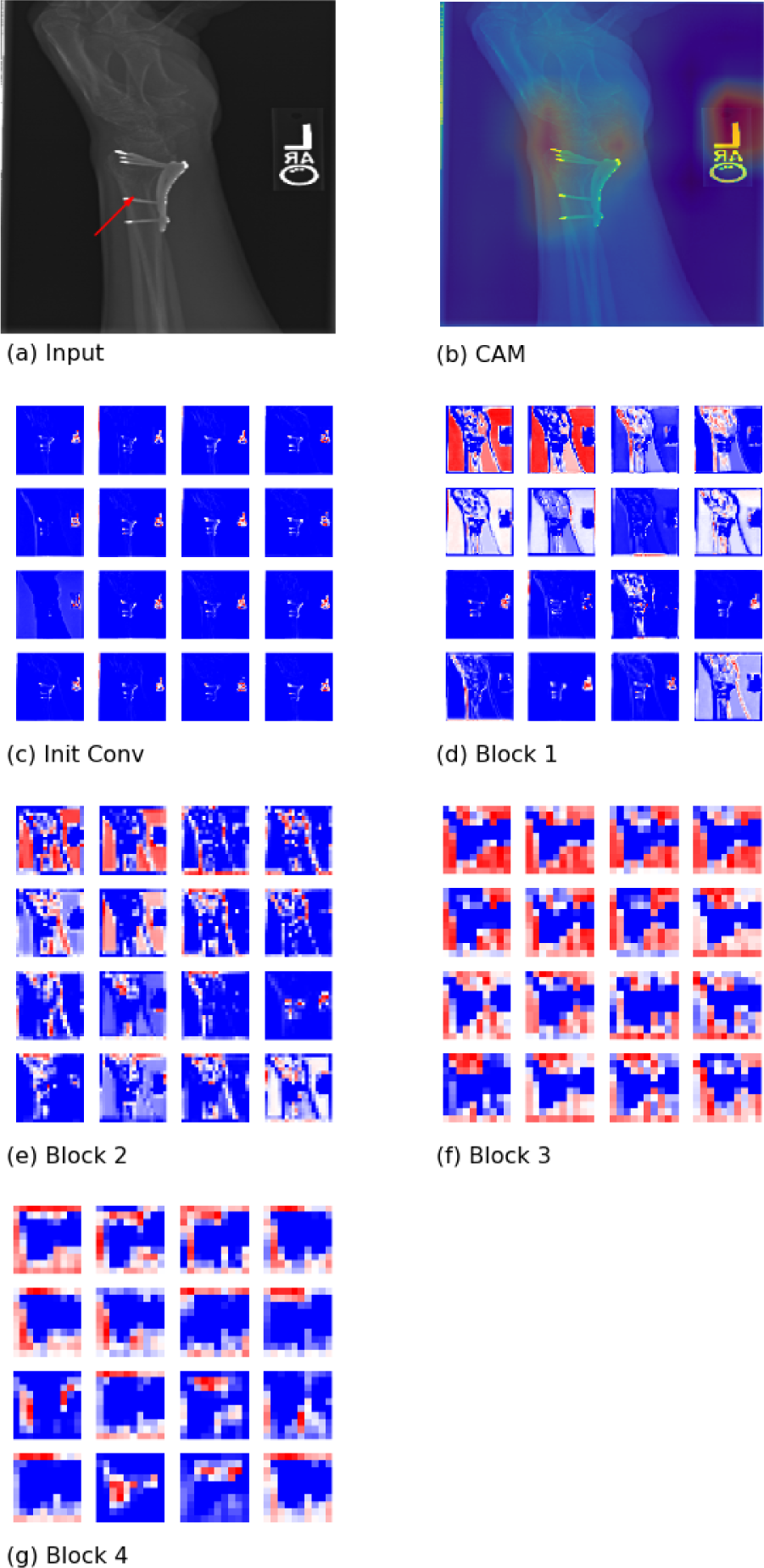
Feature extraction from an image with screw fixation at the wrist with a randomly initialised model. Poor foreground separation can be seen in (c). Only the text legend could be identified. The poor extraction reduced the capacity of the model for extracting finer features across the subsequent blocks(d - g). The final features were primarily spanning across the entire middle range of the image (g). With poor identification of the wrist, the class activation map (b) could only show the identified feature from the text legend.

**Fig. 8.**
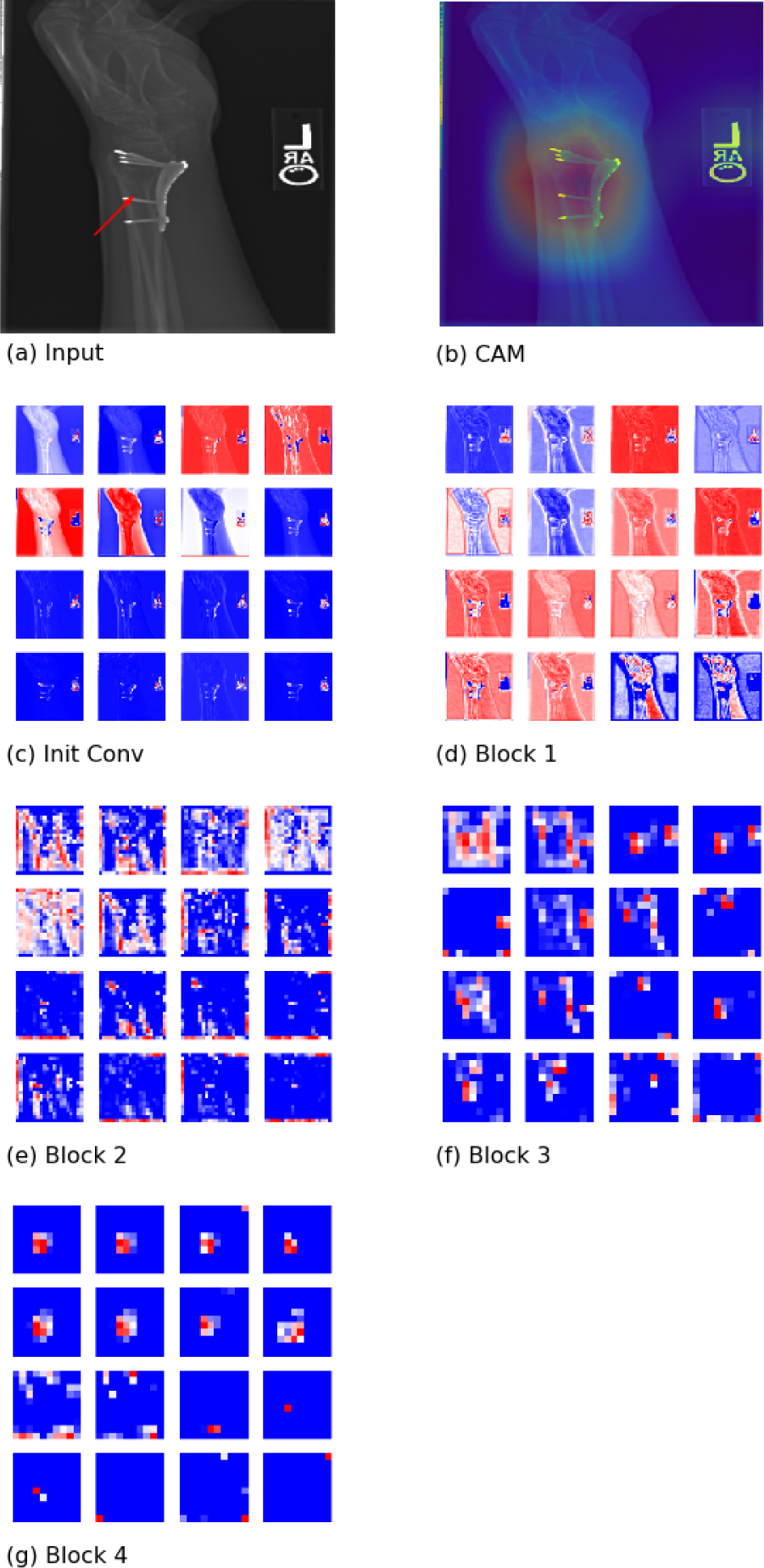
Feature extraction from an image with screw fixation at the wrist using a model trained with all extremity types. The additional trained image allowed better learning from the model to ignore the text legend as abnormality. After the foreground was identified in the initial convolution layers (c), the model tried to separate the fixation (in blue) from the text legend (in red) by the end of block 1 (d). The refinement of the key feature (screw fixation) was made from block 2 onwards (e-g). By the end of block 4, almost all the feature maps were showing only where the fixation is located (g). This had also been reflected in the class activation map (b).

**Fig. 9.**
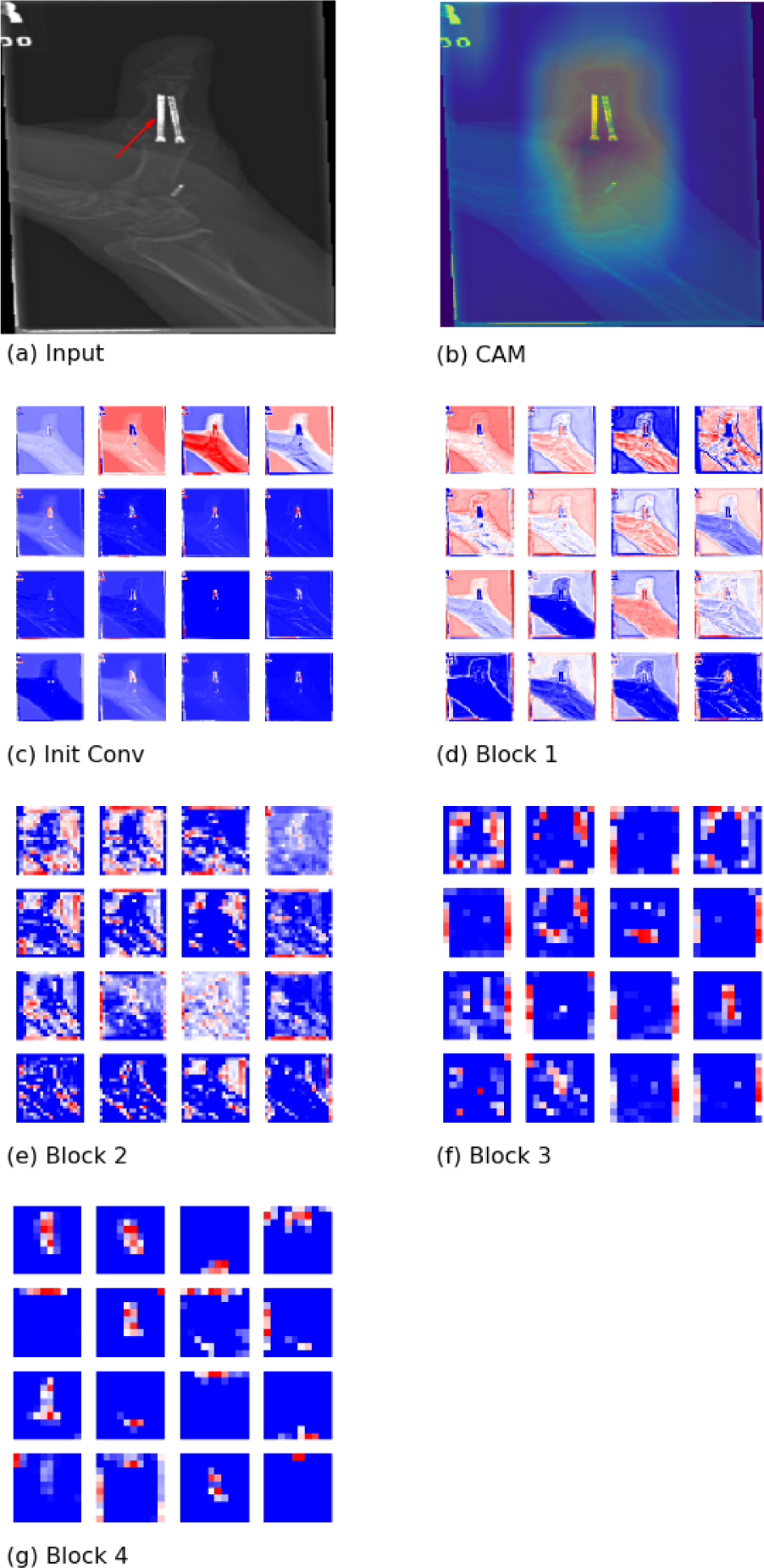
Feature extraction from an image with nails implanted in a thumb. The nails could be easily detected in a finger-only model by the end of block 1 (c & d). Refinement of key features (nails) was observed across subsequent blocks (e-g) until the thumb was the key feature extracted at the end of block 4.

**Fig. 10.**
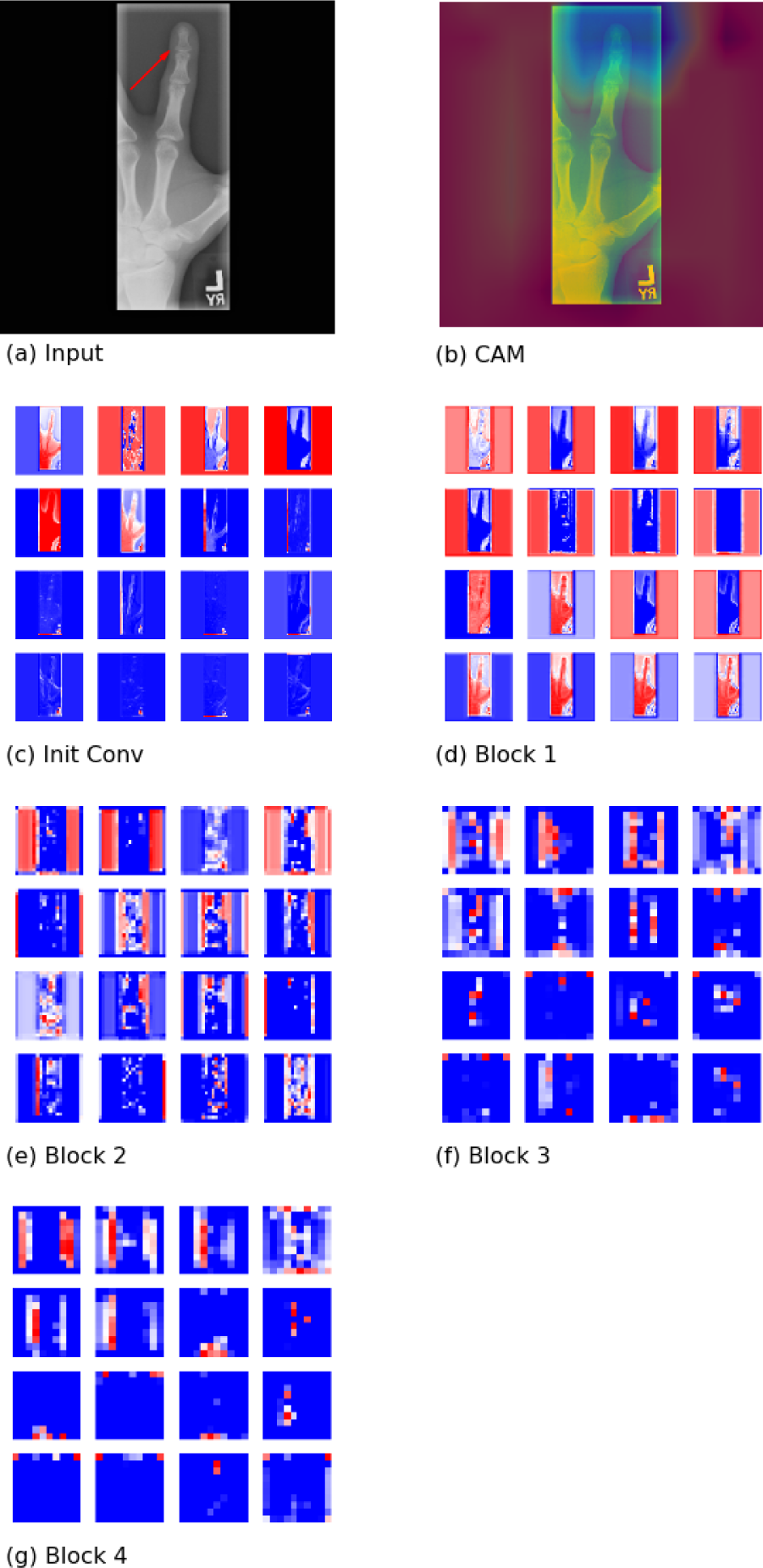
Feature extraction from an image where there is a misalignment of a fingertip. The model had taken up some capacity to separate the image frame since the initial convolution (c-g). The elimination of the frame had not been successful (e.g. blue rectangles in red background can still be seen in feature maps in block 1 & 2). This left the final attributing feature coming from the borders of the frame (g).

**Fig. 11.**
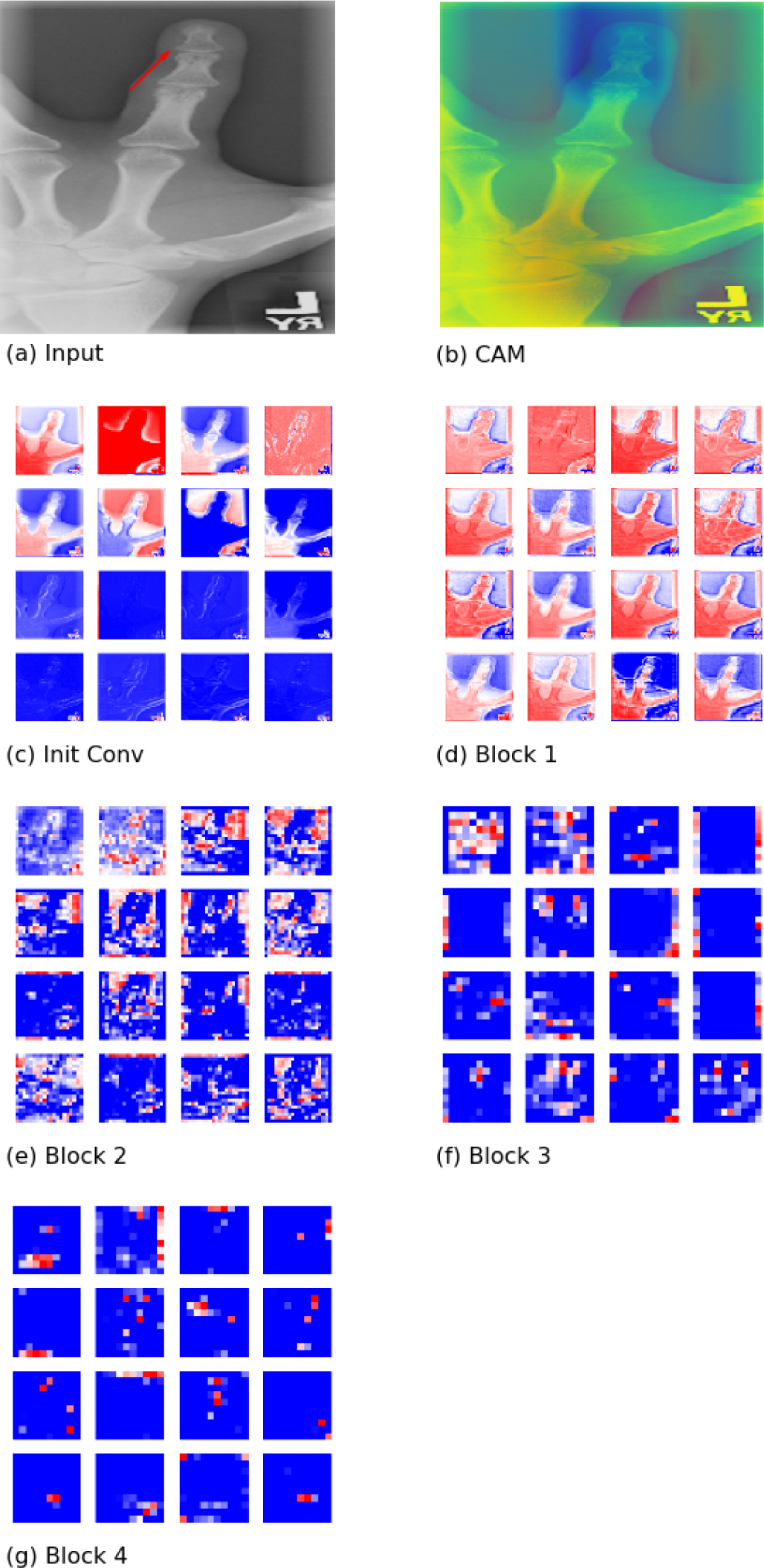
Feature extraction from a cropped image in Case 7. With the removal of the rectangular frame, the hand and finger could be identified by the end of block 1 (c & d). However, it appeared that fine-grained features (i.e. bone structures of the fingertip) remained a challenge and irrelevant features were identified across the feature maps in block 3 and 4 (f & g).

## 5. DISCUSSION

The experimental findings presented in this work have been insightful to the authors. This framework of model visualisations has enabled us with a clear representation of the feature extraction process in a trained DenseNet architecture for abnormality detection. The underlying mechanism has been found intuitive with high similarity to the approach a human would have performed. To take the example from Case 2 in Appendix A.2, the model will first try to separate the object of focus (i.e. the hand and wrist) from any background and text legends. It will then be zoomed into the trained region (i.e. wrist) for relevant features and subsequently went deeper into individual bone structures. It is worth mentioning that the actual operation for abnormality detection will require two projections of the same case from a clinical perspective.

By utilising this framework of visualisations, key information about the capability of the 169-layered DenseNet model can be revealed. For example, it can be made clear that a potential shallower network can be applied for easier detection (i.e. hardware implants) with better computational efficiency. [11] were able to obtain excellent performance in a relatively simple CIFAR-10 classification task using a DenseNet architecture of only 100 layers.

The value of using ImageNet for transfer learning as pre-trained weights has been clearly demonstrated to be crucial for better object identification. It has also proven that knowledge imparted from a source of unrelated images can still be highly beneficial for this target abnormality detection. This finding is consistent with observations made from other literature [17, 18, 13]. The issue with poor detection of degenerative joint diseases can be attributed to the lack of relevant knowledge for fine-grained feature extraction. This may be resolved by utilising more visually similar images for training [13].

This work has also presented how background noise (e.g. extra image frame) can have a long-lasting effect across the network impacting the model predictability. The model has to find the correct object of focus before any fine-grained feature can be extracted. With the use of our proposed framework, the root cause can be identified and the mitigating effect can be immediately visualised.

Finally, it has also drawn to our attention that an accurate model by statistical metrics may not actually perform as expected. Abnormality can be detected based on wrongly chosen features. This can typically be left unnoticed without the use of appropriate model visualisations. The consequence can be detrimental especially in the medical field. As such, it is recommended to use this framework to support any decisionmaking process from a predictive model with an explanation on how the outcome is generated.

## 6. CONCLUSION

The investigations carried out in this paper have created a new perspective for understanding the feature extraction mechanism within a deep convolution neural network (e.g. DenseNet). Such understanding can be achieved through the proposed framework of model visualisation using layer-wise activation and class activation maps.

This work has revealed a close resemblance of a trained DenseNet model for interpreting X-ray abnormality to the approach of a human. This resemblance should promote a more natural model explanation of the feature extraction process in complex deep learning models. Communication between deep learning model architects and medical practitioners can therefore be improved. It is expected that the framework can be applied almost directly to other networks and data sets, with potential to become a practical toolkit for future deep learning model development and application in radiology.

With the highly visual activation maps, model optimization issues otherwise hidden can be identified (e.g. a need for image pre-processing or further model training/fine-tuning). At the same time, the features extracted from an optimized model will allow medical practitioners to focus on specific regions of analysis with high confidence, as domains experts has indicated. This will streamline workflow e.g. in hospital A&Es under strict time constraint.

This work is being extended to model interpretation and visualization in other use cases and will be evaluated extensively next in the context of radiology in collaboration with hospital experts. The incorporation of attention [19] to help interpretation of areas of interest is a promising next step in the research. In many applications of deep learning, interpretation and explanation techniques will accelerate the development and deployment of deep learning architectures which can be trained with high precision and trusted at accomplishing a target task.

## Data Availability

All data used in this paper is collected from the publicly available MURA dataset (https://stanfordmlgroup.github.io/competitions/mura/) with permitted use from the data maintainer.
No other data has been made available.

### A. APPENDIX

#### A.1. Distribution & Leaderboard for the MURA Dataset

#### A.2. Supplementary activation mappings

*A.2.1. Case 2: Case of Screw Fixation at Wrist using a Pre-Trained Wrist-Only Model*

*A.2.2. Case 3: Case of Degenerative Wrist Joint Disease using a Pre-Trained Wrist-Only Model (False Negative)*

*A.2.3. Case 4: Case of Screw Fixation at Wrist using a Random Initialised Wrist-Only Model*

*A.2.4. Case 5: Case of Screw Fixation at Wrist using a Pre-Trained All Extremity Model*

*A.2.5. Case 6: Case of Nails in the Thumb using a Pre-Trained Finger-Only Model*

*A.2.6. Case 7: Case of Misaligned Fingertip using a Pre-Trained Finger-Only Model*

*A.2.7. Case 8: Case of a Cropped Version of the Misaligned Fingertip using a Pre-Trained All Extremity Model*

